# Theta Burst Stimulation in Patients With Methamphetamine Use Disorder: A Meta-Analysis and Systematic Review

**DOI:** 10.1101/2025.03.24.25324326

**Authors:** Gopalkumar Rakesh, Thomas G. Adams, Dylan H. Ballard, Christopher J. McLouth, Craig R. Rush

## Abstract

Novel interventions are urgently needed to treat methamphetamine use disorder (MUD), for which there are no FDA-approved treatments. Previous studies in patients with MUD suggest transcranial magnetic stimulation (TMS) over the left dorsolateral prefrontal cortex (L. dlPFC) decreases craving for methamphetamine. Theta burst stimulation (TBS), which includes intermittent TBS and continuous TBS (cTBS), is increasingly being used for substance use disorders, including MUD. Previous reviews of TMS in MUD performed sub-group meta-analyses of studies that delivered TBS in MUD. However, these meta-analyses included studies with overlapping participant cohorts. Given the absence of prior meta-analyses or reviews examining TBS in MUD using unique participant cohorts, we reviewed randomized controlled trials (RCTs) from three databases (PubMed/Medline, EMBASE, Google Scholar) until September 1, 2024, comparing the impact of TBS versus sham TBS on cue-induced methamphetamine cravings in patients with MUD. We performed a meta-analysis with four eligible RCTs that delivered iTBS. Results suggest iTBS was more effective in reducing cue-induced methamphetamine cravings than sham iTBS (standardized mean difference [SMD] in change = 1.04; 95% CI [0.16, 1.92]). Our systematic review included two additional RCTs that did not have sham comparator arms; one of these demonstrated a significant reduction in methamphetamine craving with accelerated iTBS. Future studies should examine if iTBS can impact clinical outcome measures other than craving, such as methamphetamine use, by measuring return to drug use. It is also pertinent to explore accelerated iTBS and cTBS for MUD and study their effects on relevant biomarkers for MUD.

## 1. Introduction

### 1.1. Background

The United States is in the grip of an unprecedented drug abuse and overdose crisis (1). Psychostimulant use, primarily methamphetamine, was the second largest cause of death from overdose (34,855) in 2023 (2). Methamphetamine use disorder (MUD) is associated with harmful consequences, including cardiovascular events (3), increased transmission of HIV and hepatitis C (4, 5), cognitive deficits (6), poor dental health (7), and higher rates of comorbid tobacco and cannabis use disorders compared to the general population (8, 9).

Currently, no FDA-approved treatments exist for MUD (10). Medications examined for efficacy in MUD include antidepressants (bupropion, mirtazapine, sertraline, atomoxetine), antipsychotics (aripiprazole), psychostimulants (modafinil, dexamphetamine, methylphenidate), anticonvulsants (topiramate), muscle relaxants (baclofen) and medications used for other substance use disorders (e.g., naltrexone, varenicline, and buprenorphine) (10, 11). Only bupropion (12), naltrexone (13–16), mirtazapine (17, 18), and methylphenidate (19, 20) showed some benefit for MUD. Medication combinations tested for MUD include: 1) naltrexone and alprazolam (21, 22); 2) bupropion and naltrexone (23–25); 3) naltrexone and N-acetylcysteine (26); and 4) flumazenil, gabapentin, and hydroxyzine (27, 28). The difficulty identifying a widely effective pharmacotherapeutic and the epidemiological data noted above underscores the urgent need to identify novel interventions for MUD.

Transcranial magnetic stimulation (TMS) is a method of noninvasive neuromodulation with demonstrated efficacy in the treatment of several substance use disorders (29). The effects of TMS on the brain depend on several factors, but a few are particularly salient. In addition to the brain state during stimulation (30), TMS parameters affecting the brain include frequency (e.g., high vs. low), number of pulses delivered, stimulation intensity, intervals between pulses, coil type (e.g., figure-8 vs. deep TMS coils), number of sessions (single vs. multiple), accelerated vs. non-accelerated (multiple sessions/day vs. a single session/day), and brain region(s) targeted (31). Theta burst stimulation (TBS) is a unique TMS paradigm with theta frequency stimulation nested within gamma frequency to deliver significantly more pulse doses within shorter periods than other repetitive TMS paradigms (32). TBS is of 2 types – intermittent TBS (iTBS), which has gaps between stimulus trains and is excitatory, and continuous TBS (cTBS), which does not have gaps and is inhibitory (33).

### 1.2. Current evidence

Deep TMS received FDA approval as a short-term smoking cessation aid in 2021 (34). A systematic review of 51 studies examined the effects of TMS in people with substance use disorders (29). Sub-group meta-analyses revealed that TMS reduced craving for alcohol and tobacco with significant standardized mean differences (SMD) (0.86-1.65) when multiple daily TMS sessions targeted the left dorsolateral prefrontal cortex (L.dlPFC) (29). The authors conducted a qualitative systematic review of all TMS studies in MUD but did not perform a meta-analysis of TBS for MUD (29).

Previous systematic reviews and meta-analyses of TMS studies (including TBS) in patients with MUD have shown preliminary evidence of TMS reducing cravings for methamphetamines (35–40). Two of these reviews were scoping (38) and narrative (39) in their formats, focusing on all TMS paradigms in MUD. One review of TMS studies (including TBS) in MUD performed a meta-analysis showing a large SMD (Hedge’s g = 1.54, 95% CI: 0.75-2.35) for TMS compared to sham for methamphetamine craving (35). Two previous meta-analyses of all TMS studies in MUD conducted sub-group meta-analyses of iTBS; they showed large SMDs of 1.22 (95% CI: 0.95–1.48, P < 0.001, 4 studies) (36) and 1.08 (95% CI: 0.81-1.36, P<0.001, 5 studies) (37) for iTBS compared to sham iTBS.

However, these sub-group meta-analyses were disadvantaged in including multiple MUD studies with overlapping participant cohorts (36, 37). This limited the accuracy and interpretation of results. In addition, neither meta-analysis included a notable RCT comparing the effects of 1800 pulses of iTBS to sham iTBS on methamphetamine craving (41). Lastly, a previous meta-analysis examined the dose-response curve for change in desire for drug questionnaire (DDQ) in MUD with iTBS (40).

### 1.3. Study objective

Given the increasing relevance of TBS for substance use disorders (29) and MUD (38), we reviewed unique RCTs with non-overlapping participant cohorts with MUD to perform a meta-analysis examining the effect of TBS (iTBS and cTBS) relative to sham TBS on cue-induced methamphetamine craving. We also conducted a qualitative systematic review of TBS studies in MUD.

## Methods

The protocol for the review and meta-analysis was developed and registered a priori (PROSPERO ID: CRD42024590977). We followed the Preferred Reporting of Items for Systematic Reviews and Meta-Analyses (PRISMA) guidelines for reporting the purpose, methods, and results of this meta-analysis and systematic review (42).

### 1.4. Data sources and searches

We searched three databases (PubMed/Medline, Google Scholar, and EMBASE) using the terms ‘theta’, ‘burst’, ‘stimulation’, and ‘methamphetamine’ for studies that examined the effects of theta burst stimulation in people with MUD, published in English from inception till September 1, 2024.

### 1.5. Study selection

Before deciding to include or exclude the study in the meta-analysis, two authors (GR and DB) screened them by title, abstract, and full text. Studies to include in the meta-analysis were screened for the following inclusion/exclusion criteria: 1) the study needed to be an RCT; 2) participants in the study met the criteria for MUD as defined by any edition of the Diagnostic Statistical Manual of Disorders (DSM) or International Classification of Diseases (ICD); 3) administered either iTBS or cTBS with sham comparator; 4) cue-induced methamphetamine craving was an outcome measure in the study, measured with the visual analog scale (VAS); 5) the study involved adult participants ≥ 18 years of age; and 6) the RCT had a unique clinical trial identification number to avoid overlap of participant cohorts between RCTs. We examined the full texts of all candidate studies using the clinical trial identification numbers to assess for unique non-overlapping participant cohorts. For studies with overlapping participant cohorts, we included only those that analyzed data from the largest number of participants.

Studies considered for inclusion in the qualitative systematic review met all inclusion and exclusion criteria as the meta-analysis, except that they did not need a sham comparator arm.

### 1.6. Outcome measure

The primary outcome measure in our meta-analysis was cue-induced methamphetamine craving, as measured by the VAS.

### 1.7. Data extraction

We (GR and CM) extracted the following data from all studies included in the meta-analysis - craving scores (average and standard deviation) and changes in craving scores (average and standard deviation) when available. We extracted the following additional facets for studies included in the systematic review – participant sample sizes, number of study arms, TBS parameters including type of TBS (iTBS versus cTBS), number of pulses, stimulation threshold, and number of sessions.

### 1.8. Data synthesis and analyses

A standardized mean difference (SMD) in change from pre-to post-treatment was used to measure the effect size of interest using a pooled standard deviation (SD) with heteroscedastic population variances. The meta-analysis used a random effects model via restricted maximum likelihood (REML) to account for the potential heterogeneity between studies. Pooled SMDs and associated 95% confidence intervals (CI) were calculated using the Knapp and Hartung adjustment (43). A heterogeneity (*I^2^*) measure was calculated, and a funnel plot was examined. The funnel plot was generated by plotting the observed effect sizes or outcomes on the x-axis against the corresponding standard errors on the y-axis. For all studies included in the meta-analysis, the change scores’ SD was calculated using a correlation coefficient of 0.6, using the method elucidated in the Cochrane Handbook for Systematic Reviews of Interventions (44). We then performed a sensitivity analysis using two other correlation values (0 and 0.5) to calculate the SD of the change scores (45). All analyses were performed using Microsoft Excel and R Statistical Software (v4.1.3; R Core Team) using the metafor package (46).

## 2. Results

### 2.1. Description of studies included in the meta-analysis

Figure 1 shows the PRISMA diagram of study selection for the meta-analysis and the systematic review. We performed the meta-analysis with four studies that compared the effects of iTBS versus sham iTBS on methamphetamine craving (41, 47–49). All four studies used unique non-overlapping participant cohorts, as identified from their clinical trial identification numbers (41, 47–49). The results of the random effects meta-analysis suggest iTBS was more effective in reducing cue-induced methamphetamine cravings than sham iTBS (standardized mean difference in change = 1.04; 95% CI [0.16, 1.92]) (Figure 2). The studies had high heterogeneity (*I^2^* = 67%, p = 0.02). Figure 3 displays an asymmetrical funnel plot, indicating the presence of publication bias. Sensitivity analyses using 0 and 0.5 as correlation coefficients to calculate the change’s SD resulted in standardized mean differences of 0.74 (95% CI [0.24, 1.25]) and 0.96 (95% CI [0.17, 1.74]).

**Figure 1.**
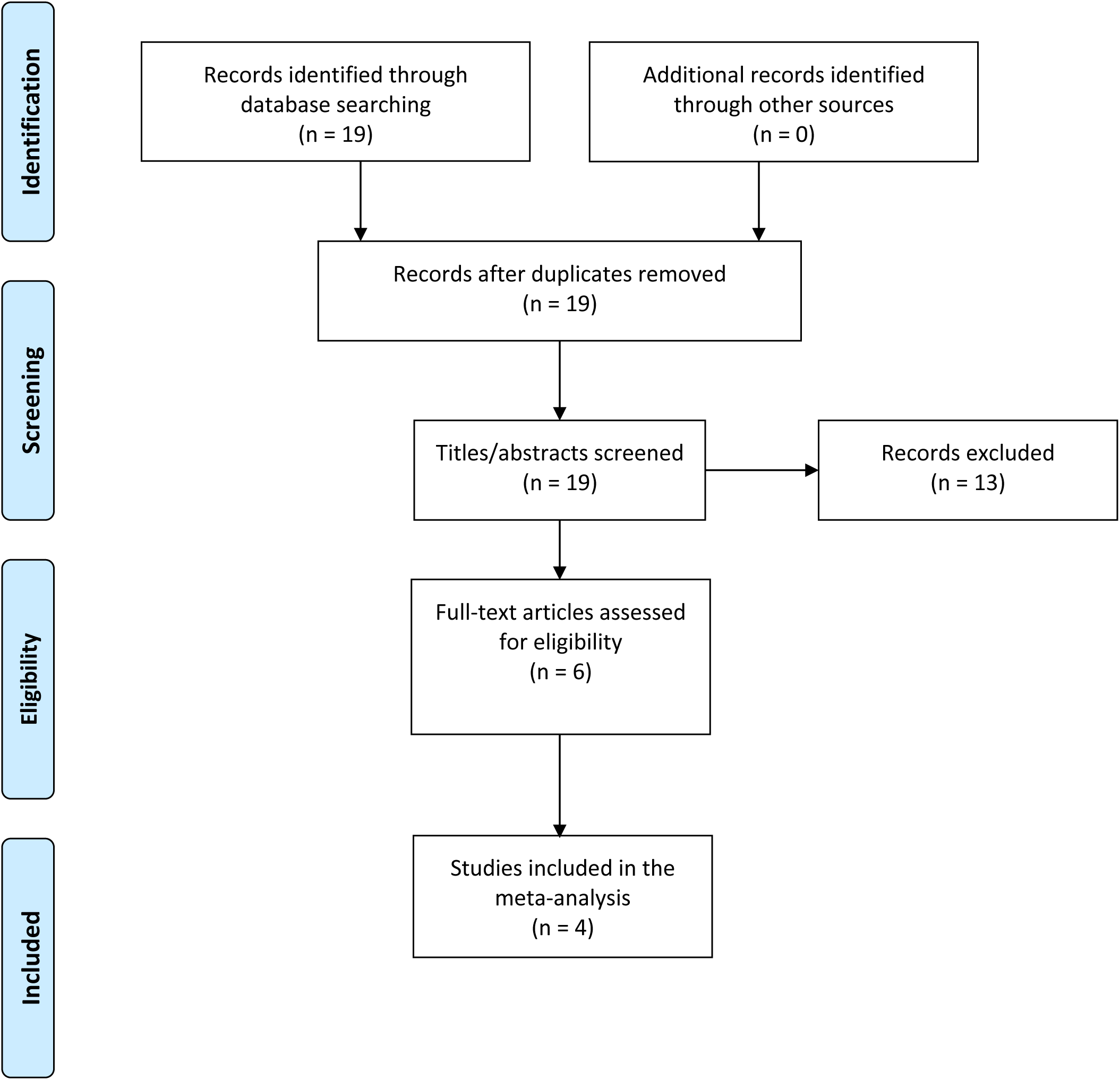
PRISMA diagram of the study selection process. The PRISMA diagram shows how selected studies were included in the review and meta-analyses. PubMed/Medline, Google Scholar, and EMBASE were the databases screened for studies. All studies were published in English and screened by title, abstract, and full text by two authors (GR and DB) before deciding whether to include or exclude them.

**Figure 2.**
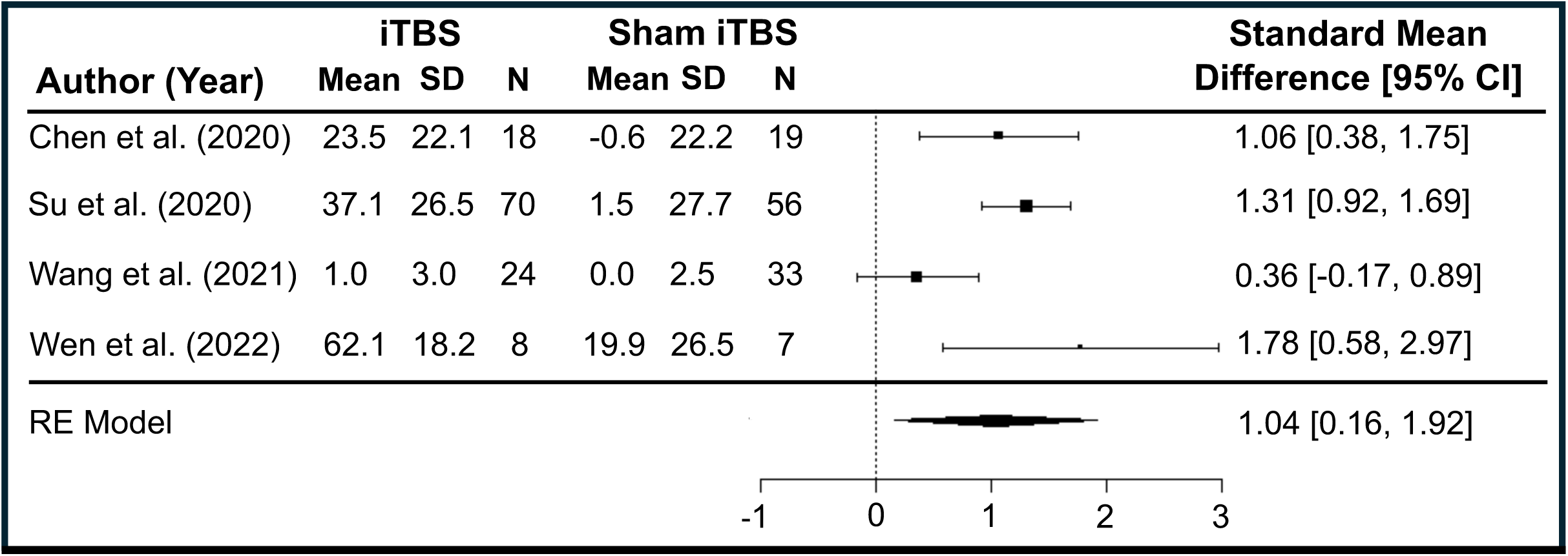
Forest Plot of Standardized Mean Differences. This is a forest plot of the random effects meta-analysis demonstrating that iTBS was more effective in reducing cue-induced methamphetamine cravings than sham iTBS (standardized mean difference in change = 1.04; 95% CI [0.16, 1.92]). The studies had high heterogeneity (*I^2^* = 67%, p = 0.02).

**Figure 3.**
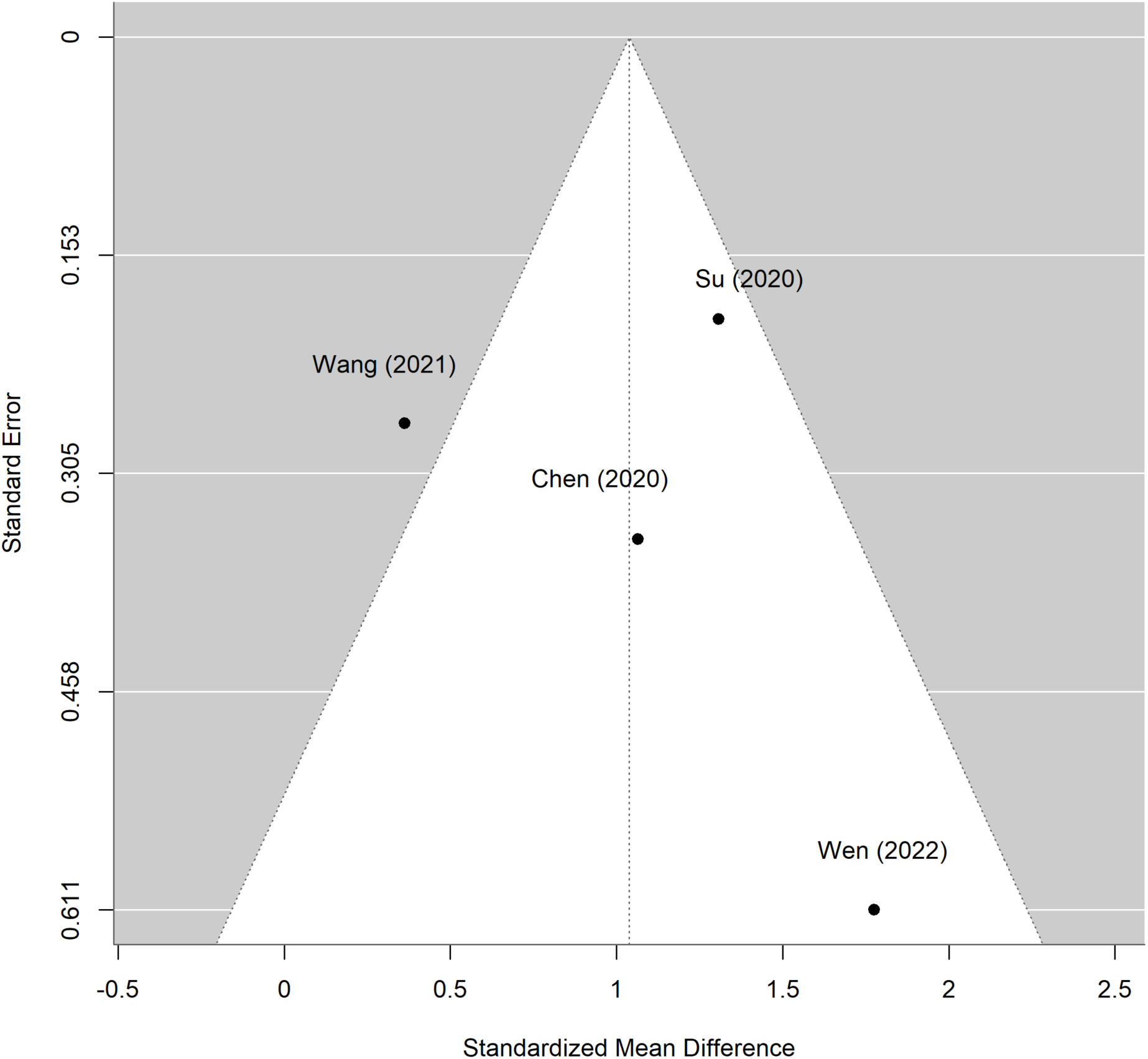
Funnel Plot of Standard Error by Standardized Mean Difference The funnel plot was generated by comparing the observed effect sizes or outcomes on the x-axis to the corresponding standard errors on the y-axis. One study (49) was outside the funnel plot, indicating publication bias.

The L.dlPFC was the most common stimulation site across the four studies. The number of pulses per session ranged from 600 (49) to 900 (47, 48) and 1,800 (41). The number of sessions varied across trials, ranging from 2 to 20. Wen et al. delivered only two sessions of iTBS (1,800 pulses/session) but showed a higher effect size (1.78) than the other three trials, which delivered 20 sessions but a lower dose (600 to 900 pulses per session) (41, 47–49).

Only one study compared cTBS to sham TBS (48). This study compared the effects of combining iTBS and cTBS on methamphetamine craving versus delivering them separately or delivering sham TBS (48). The iTBS was delivered over L. dlPFC, and cTBS over the ventromedial prefrontal cortex (vmPFC) (48). The combination of iTBS (L.dlPFC, 900 pulses) and cTBS (vmPFC, 900 pulses) showed the most significant reduction in cue-induced methamphetamine craving compared to the other three arms 1) 900 iTBS pulses to the L. dlPFC, 2) 900 cTBS pulses to the vmPFC, and 3) sham TBS (48).

### 2.2. Description of studies included in the systematic review

The systematic review encompassed the four RCTs included in the meta-analysis (41, 47–49) and two additional RCTs that did not have sham comparator arms (50, 51) (Table 1). Table 1 lists these six studies. Among the two additional RCTs in the systematic review, one RCT delivered accelerated iTBS in MUD (51). This study delivered two sessions daily (separated by four hours) for five days and compared three arms: 1) 600 iTBS pulses to the L.dlPFC, 2) 600 cTBS pulses to the L.dlPFC, 3) 600 cTBS pulses to the right dlPFC (51). Cue-induced methamphetamine craving decreased significantly in the iTBS-L.dlPFC group and cTBS-right.dlPFC group, but not in the cTBS-L.dlPFC group (51). The other RCT compared 600 pulses iTBS to 10 Hz TMS. Both paradigms decreased cue-induced methamphetamine craving when given daily for 12 days, but there was no significant group by time interaction (50).

**Table 1.**
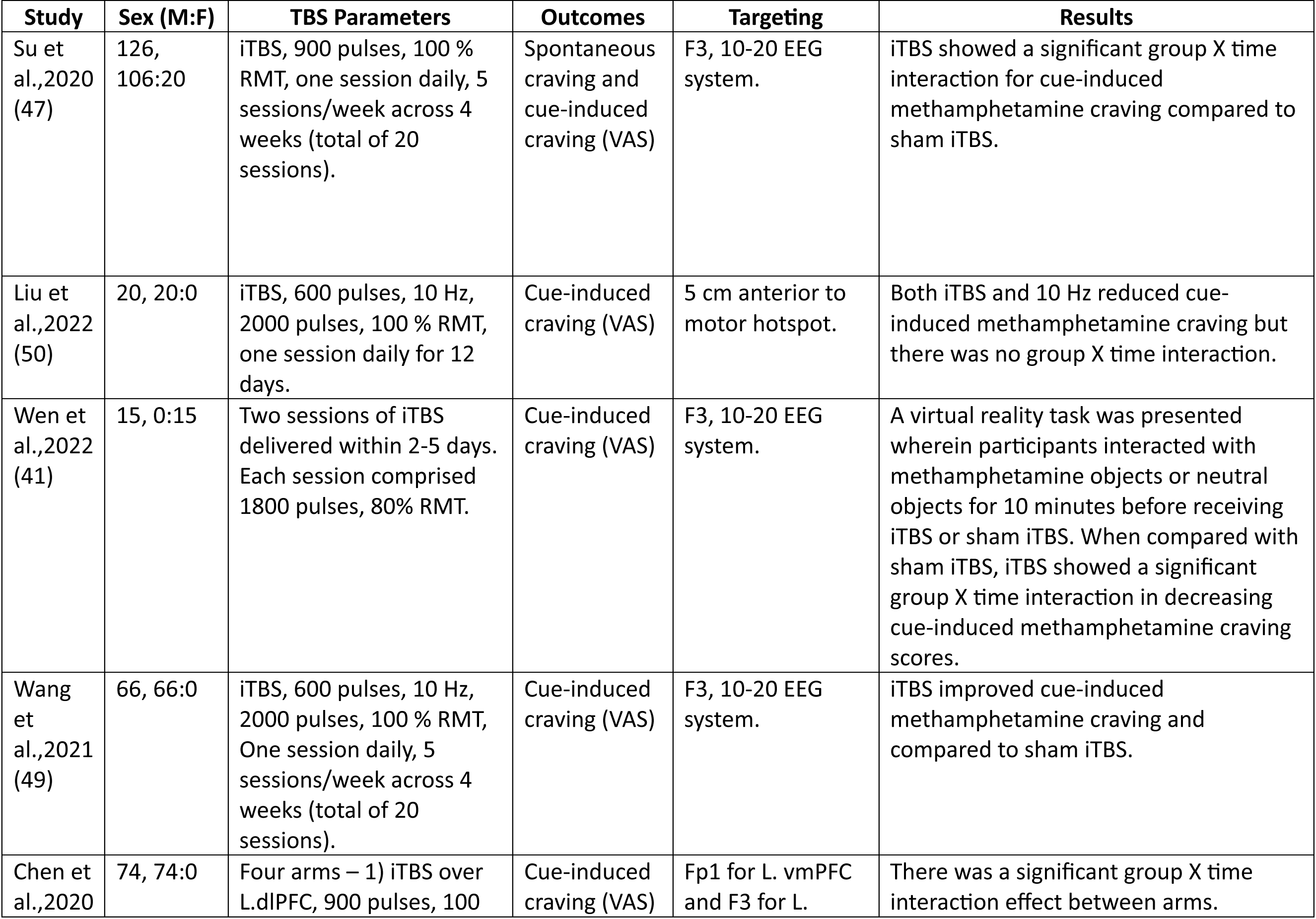
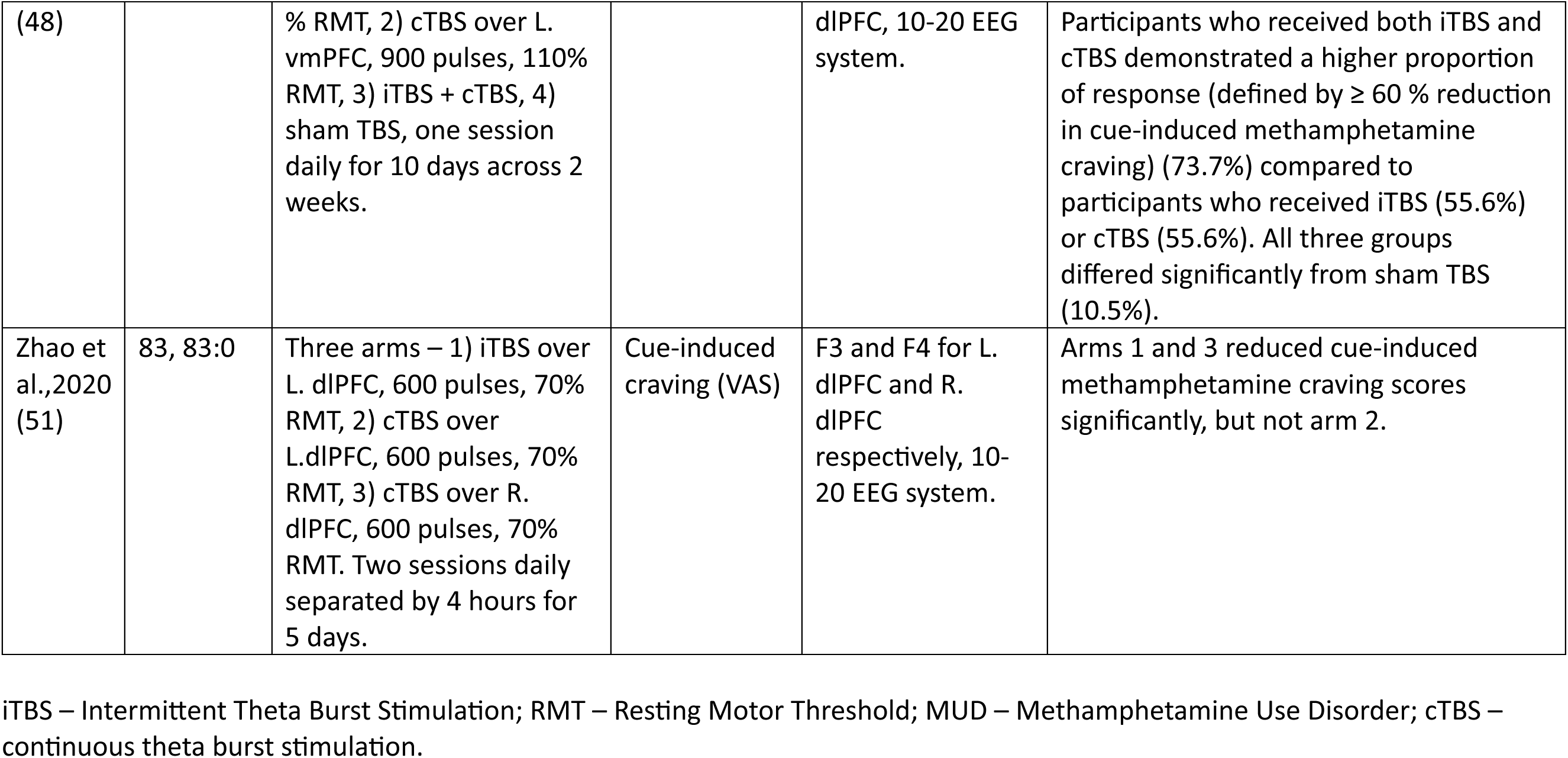
Summary of Included Studies in the Systematic Review.

## 3. Discussion

This meta-analysis showed iTBS has greater efficacy in reducing cue-induced methamphetamine craving reduction than sham iTBS, using RCTs done to date that compared iTBS to sham iTBS in unique participant cohorts. We found only one study that compared cTBS to sham TBS and did not perform a meta-analysis to examine the impact of cTBS on methamphetamine craving.

### 3.1. iTBS as a treatment option for MUD

iTBS differs from other TMS frequency paradigms in that it is shorter in duration and more efficient (32). This helps with treatment tolerability and patient retention in substance use disorders (29). In MUD, iTBS doses ranged from 600-1,800 pulses/session and duration from 2 to 20 days. Although Wen et al. delivered only two sessions of iTBS, they showed a higher magnitude of difference in craving scores between iTBS and sham iTBS than the other three trials that delivered 20 sessions (41, 47–49). This could be due to the higher number of pulses (1,800) delivered per session by Wen et al. compared to the other three trials that delivered 600-900 pulses per session. Nonetheless, the heterogeneity in iTBS frequency parameters and doses across RCTs precludes any inferences on the impact of pulse doses or the number of sessions on craving. This also makes it challenging to generate a consensus on a future dosing regimen or treatment protocol to address clinical outcomes such as methamphetamine use in MUD, quantified with urine drug screens or self-reports.

Protocols that deliver more than one session daily are called accelerated protocols (52, 53). One study delivered accelerated iTBS in MUD, consisting of two sessions daily for 5 days (51). The accelerated iTBS study in MUD did not have a sham comparator arm (51). Nonetheless, there was a reduction in craving in the iTBS arm (51). Progression toward an accelerated iTBS protocol for MUD could incorporate more sessions daily, building on the Stanford Neuromodulation Treatment (SNT) protocol, which is FDA-approved for treatment-resistant major depressive disorder (MDD) (54). The SNT protocol delivered ten sessions of iTBS (1,800 pulses per session) daily for five days, with 50-minute intervals between sessions in patients with treatment-resistant MDD (54). SNT uses external hardware and the patient’s resting state functional connectivity (rsFC) between the subgenual cingulate and L. dlPFC (54) to localize the iTBS target and individualize the TMS target (55).

Translating the SNT protocol to MUD would require parametric dosimetry studies that carefully consider seizure risk in MUD and plateau effects from total stimulation doses (40, 56, 57). Higher dose delivery may not lead to a further reduction in craving, as demonstrated by a dose-response meta-analysis of RCTs that used iTBS in MUD (40). With the optimal dose number unique to diseases, MUD had a plateau in dose response at 9,724 pulses, as assessed using the DDQ (40). However, there was considerable heterogeneity (∼70%) in studies utilized for this analysis (40). Designing an accelerated iTBS protocol for MUD would also need to encompass precision targeting using neuroimaging.

### 3.2. Utilizing neuroimaging to target TBS in MUD

Regardless of the stimulation site, delivering TMS to a cortical target influences neural activity at other brain regions connected structurally or functionally with the stimulation site (58, 59). Thus, TMS directed at a specific brain network could also lead to downstream effects at other brain networks besides the index one.

While there has been debate regarding the need for targeting with neuroimaging, recent evidence suggests more significant clinical benefits in MDD when delivering TMS with resting state functional connectivity (rsFC) based targeting than scalp-based targeting techniques (60, 61). There is a consistent rsFC signature predicting TMS response in MDD (61). The efficacy of TMS is positively associated with the rsFC anticorrelation between the subgenual anterior cingulate and the L. dlPFC (62, 63) and the distance between the final stimulation site and the personalized L. dlPFC target (55, 64).

Unlike MDD, we do not have a consistently replicated rsFC-based TMS target in MUD. However, research suggests several promising options for investigation. Summarizing the current landscape of studies examining rsFC in patients with MUD makes it pertinent to discuss the Impaired Response Inhibition Salience Attribution (iRISA) model (65). This model encompasses network dysfunction in the three higher-order brain networks (salience network [SN], central executive network [CEN], and default mode network [DMN]) in addition to the reward network (65). Based on this model, previous rsFC studies in stimulant use disorders (encompassing cocaine, amphetamine, methamphetamine, and combinations thereof) showed decreased rsFC in the following network pairs - 1) within CEN and SN, 2) between CEN and DMN, 3) between reward network and DMN, and 4) between SN and DMN, and increased rsFC in the following network pairs – 1) between CEN and reward network, and 2) between CEN and SN (66). These network abnormalities are reliably observed regardless of other illness facets (type of stimulant used, abstinence versus ongoing use, years of use) (66).

Four studies examined rsFC specifically in MUD without any interventions (67–70). One was notable in showing increased rsFC within the DMN and between the DMN and the CEN compared to healthy controls, both of which correlated with methamphetamine use (67). Compared with sham iTBS, delivering 20 iTBS sessions (900 pulses/session) over 4 weeks (1 session/day, 5 sessions/week) increased rsFC between the L.dlPFC and the left inferior parietal lobule (which was negatively correlated with change in methamphetamine craving). iTBS also decreased rsFC between the right anterior insula and the left medial temporal gyrus, the left anterior insula and the right inferior parietal lobule/left superior parietal lobule, and the right anterior insula and the right inferior parietal lobule/right angular gyrus (71). Considering the iRISA network model, these results are consistent with decreased rsFC between DMN (left medial temporal gyrus) and SN (right anterior insula), CEN (right inferior parietal lobule/left superior parietal lobule) and SN (left anterior insula), DMN (right angular gyrus) and SN (right anterior insula).

Most TBS studies in MUD targeted the L.dlPFC (Table 1), part of the central executive network (72, 73). There is potential value in exploring an rsFC signature in MUD based on the iRISA model as a biomarker of response to TBS. In addition, a suitable avenue to explore would be the value of fMRI drug cue reactivity (FDCR) for personalized TBS targeting to the L.dlPFC (74, 75). Although no previous study used FDCR to target TMS in substance use disorders, this direction has been consistently recommended as a method to personalize targeting in substance use disorders (75, 76). FDCR studies across substances have demonstrated neural cue reactivity in response to the contrast between substance and neutral cues to be correlated with craving, relapse, treatment response, and long-term prognosis (75, 76). Previous FDCR studies in MUD have shown the mPFC, ventral striatum, and amygdala are consistently activated when viewing MA versus neutral cues (77–79). The consistency in results across MUD studies bolsters the argument that FDCR is more valuable for targeting TBS than rsFC-based targeting.

### 3.3 Alternative stimulation sites

The frontal pole could offer an alternative site for TBS in patients with MUD, as displayed by a previous study that demonstrated methamphetamine craving reduction with cTBS over the frontal pole (48). Mechanistic results from lesion network mapping studies and cue reactivity studies highlight the importance of the frontal pole in substance use disorders (80, 81).

Previous studies have demonstrated changes in vmPFC activity by targeting the frontal pole with TMS (82, 83). Task-based functional MRI (fMRI) studies have shown the vmPFC to be instrumental for cue reactivity in MUD (84, 85). Targeting the vmPFC using cTBS decreased brain reactivity to alcohol cues and cocaine cues compared to sham (82, 83). This reveals the potential to modulate cue reactivity to methamphetamine cues with cTBS in MUD. Future studies would need to compare changes in methamphetamine craving between iTBS over L.dlPFC and cTBS over the frontal pole.

The insula could also offer a potential alternative site for TBS in patients with MUD (86). Both the anterior and posterior insula regulate interoception and decision-making, making them critical to the regulation of methamphetamine craving. Delivering TBS to the insula can be done in two ways: 1) stimulating a superficial cortical structure with structural or functional connectivity with the anterior or posterior insula using a figure-8 coil (87, 88), or 2) applying deep TMS with an H coil, which can stimulate deeper structures (89). Although suprathreshold figure-8 coil TMS could directly stimulate the insula, the required depth of stimulation precludes it from being ideal (90). Deep TMS is suited for stimulating deep structures like the insula, albeit with plausible concerns about losing focality (91, 92). Insula stimulation has shown value in the treatment of tobacco use disorder with deep TMS (93), leading to FDA approval for the same.

Studies targeting these alternative sites in MUD must be followed by trials comparing them to traditional L.dlPFC TBS. Therefore, potential future directions to explore in MUD would be 1) comparing TBS over the frontal pole with L.dlPFC TBS, 2) comparing L.dlPFC TBS with TBS over the insula using figure-8 coil (utilizing rsFC with a cortical site) or deep TMS, and 3) examining the value of combining stimulation paradigms, for example, combining iTBS over L.dlPFC and cTBS over the frontal pole.

## Conclusions

No RCT has examined the impact of TBS on clinical parameters of methamphetamine use, including ongoing methamphetamine use. Our meta-analysis indicates iTBS could be a potential treatment to address clinical outcome measures in MUD. Available studies make it challenging to define a consensus dosing schedule or regimen for MUD. Hence, more research is needed to optimize accelerated TBS protocols, optimize neuroimaging-based targeting, and target alternative brain regions, like the frontal pole or the insula. In addition to targeting TBS, examining FDCR and network-based rsFC as biomarkers of response to TBS is also timely. Given that the FDA approval process for TBS is much shorter than that for medications, which take an average of 10-15 years from inception to approval, it is imperative to spearhead trials of TBS in MUD to move towards FDA approval.

## CRediT authorship contribution statement

GR and CR conceived the research question and designed the review protocol. GR, and DB systematically screened studies for inclusion. GR and CM performed data extraction and analyses. All authors contributed equally to the writing and revision of the manuscript. All authors approved the final version of the manuscript.

## Data Availability

NA

## Acknowledgment

None

